# An interpretable mortality prediction model for COVID-19 patients – alternative approach

**DOI:** 10.1101/2020.06.14.20130732

**Authors:** Peter Gemmar

## Abstract

The pandemic spread of coronavirus leads to increased burden on healthcare services worldwide. Experience shows that required medical treatment can reach limits at local clinics and fast and secure clinical assessment of the disease severity becomes vital. In [1] a model is presented for predicting the mortality of COVID-19 patients from their biomarkers. Three biomarkers have been selected by ranking with a supervised Multi-tree XGBoost classifier. The prediction model is built up as a binary decision tree with depth three and achieves AUC scores of up to 97.84±0.37 and 95.06± 2.21 for training and external test data sets, resp.

In human assessment and decision making influencing parameters usually aren’t considered as sharp numbers but rather as Fuzzy terms [2], and inferencing primarily yields Fuzzy terms or continuous grades rather than binary decisions. Therefore, I examined a Sugeno-type Fuzzy classifier [3] for disease assessment and decision support. In addition, I used an artificial neural network (*SOM*, [4]) for selecting the biomarkers. Modelling and validation was done with the identical data base provided by [1]. With the complete training and test data sets, the Fuzzy prediction model achieves improved AUC scores of up to 98.59 or 95.12 The improvements with the Fuzzy classifier obviously become clear as physicians can interpret output grades to belong to positive or negative class more or less strongly. An extension of the Fuzzy model, which takes into account the trend in key features over time, provides excellent results with the training data, which, however, could not be finally verified due to the lack of suitable test data. The generation and training of the Fuzzy models was fully automatic and without additional adjustment with the help of ANFIS from Matlab©.

## 1 Introduction

In [1] the outbreak of COVID-19 pandemic causing severe health concerns and consequences for health care services worldwide has been described in a catchy way. It is stressed that the severity of cases is putting medical services under great pressure. Furthermore, the importance of distinguishing patients that require immediate medical attention is described and that there is a lack of capacity to identify cases at imminent risk of death. So far, no prognostic biomarkers have been available to estimate the patients risks.

Consequently, the research group in [1] analysed blood samples of 485 patients from the region of Wuhan, China. Then, a state of the art machine learning algorithm was used to identify the most discriminative biomarkers. Most crucial biomarkers have been revealed through optimization of a supervised XGBoost classifier [5]. Three key features have been derived: lactic dehydrogenase (LDH), lymphocytes and high-sensitivity c-reactive protein (hs-CRP). A clinically operable decision tree (*decTree*) was developed and the decision rules with the three key features and their thresholds were devised recursively by supervised learning.

There are many other possibilities for building a classification or prediction model. I tried two of them with very little effort for creation: 1st a support vector machine (*SVM*) and 2nd a Sugeno-type [3] Fuzzy classifier (*FIS*). Both classifiers are transparent in explaining a specific input transformation to a specific classification output. The classifier *SVM* delivered binary predictions at least as accurate as the classifier *recTree*. The classifier *FIS* is different from the two others (*decTree, SVM*) as its output esteems the grade about how much the input belongs to one of two classes (*positive, neagtive*) specified as patient outcome in the data samples. This may be an advantageous property when predicting the patients risk value. In the following I shall describe the development and evaluation of the Fuzzy classifier.

There are also many possibilities for feature analysis and selection. In my approach I put emphasis on finding those features that show signatures similar to the patient outcome and that are little to not correlated. Artificial neural networks of type Kohonen can be used to map the distribution of features in the feature space into 2D component planes (maps) revealing the signature of the according feature (Self Organizing Maps *SOM* [6]). These maps can be compared visually and those maps similar to the map of patient outcome can be identified for feature selection. In addition, correlation analysis about the features can be used to determine the minimum feature set covering the feature space in an efficient way, e.g. in terms of a minimum dominant set (*MDS*). The key features selected in [1] have been confirmed this way. Furthermore, two other features (Albumin, International Standard Ratio) have been proposed and than used with the *FIS* classifier in an extended analysis.

If one looks at the determined biomarker values in the data base created by [1], one will of course notice a change in the biomarkers from the day of admission to the discharge from the hospital. It is therefore obvious to consider the trend of the biomarkers over time to the last value in the risk assessment. This is successfully examined here with an expansion of the Fuzzy model.

## 2 Data resources

Basically, all data for feature analysis as well as for training and testing the classifiers (*SVM, FIS*) have been taken from the original data base provided by [1]. This data base contains two files with sample data taken from patients: 1) *time_series_375_prerpocess_en*.*xlsx* (in the following: *train_data*) and 2) external test data *time_series_test_110_preprocess_en*.*xlsx* (in the following: *test_data*). *train_data* collects 74 biomarkers (features) together with age, gender, data sample time, admission time, discharge time, and class of patient outcome (alive, deceased) for 375 patients. *test_data* collects three biomarkers (key features) together with data sample time, admission time, discharge time, and class of patient outcome for 110 patients.

In [1] only data of the final feature samples per patient is used for training and testing of the rule decision classifier *decTree*. There are also some patients with incomplete measurements for at least one of the selected key features leaving 351 patients in data base *train_data*, and 110 patients in data base *test_data*. The distribution of patient outcome (classes alive *≡negative*, deceased*≡ positive*) over the key feature space are depicted in Figure 1 and Figure 2, resp.

**Fig. 1.**
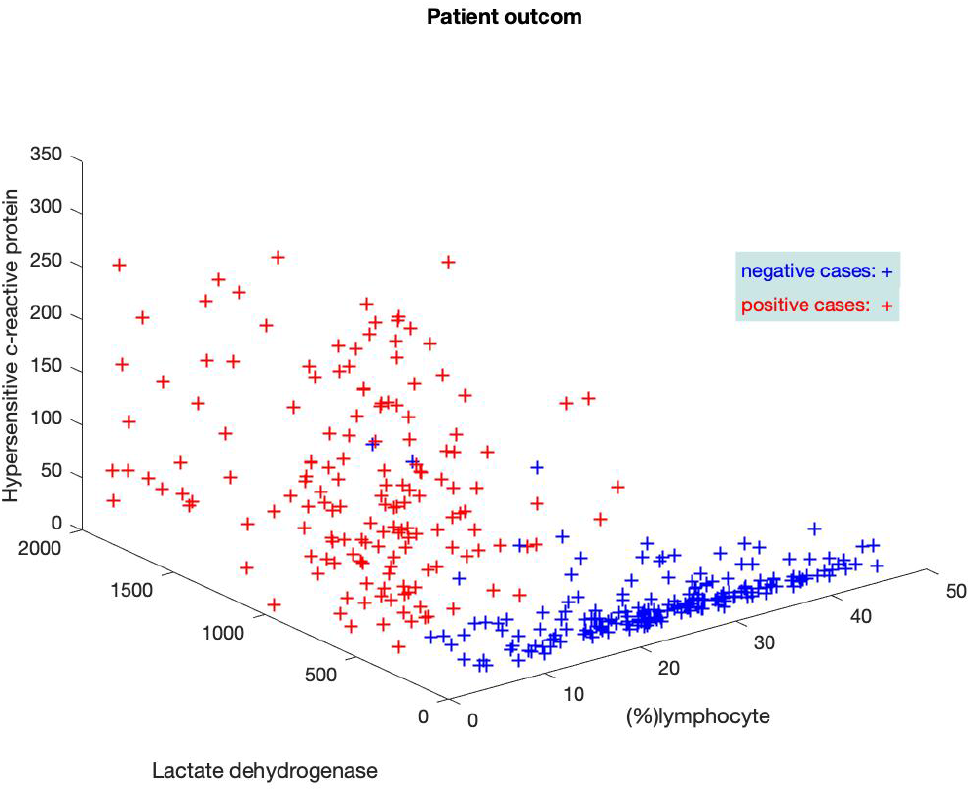
Patient outcome in *train_set*

**Fig. 2.**
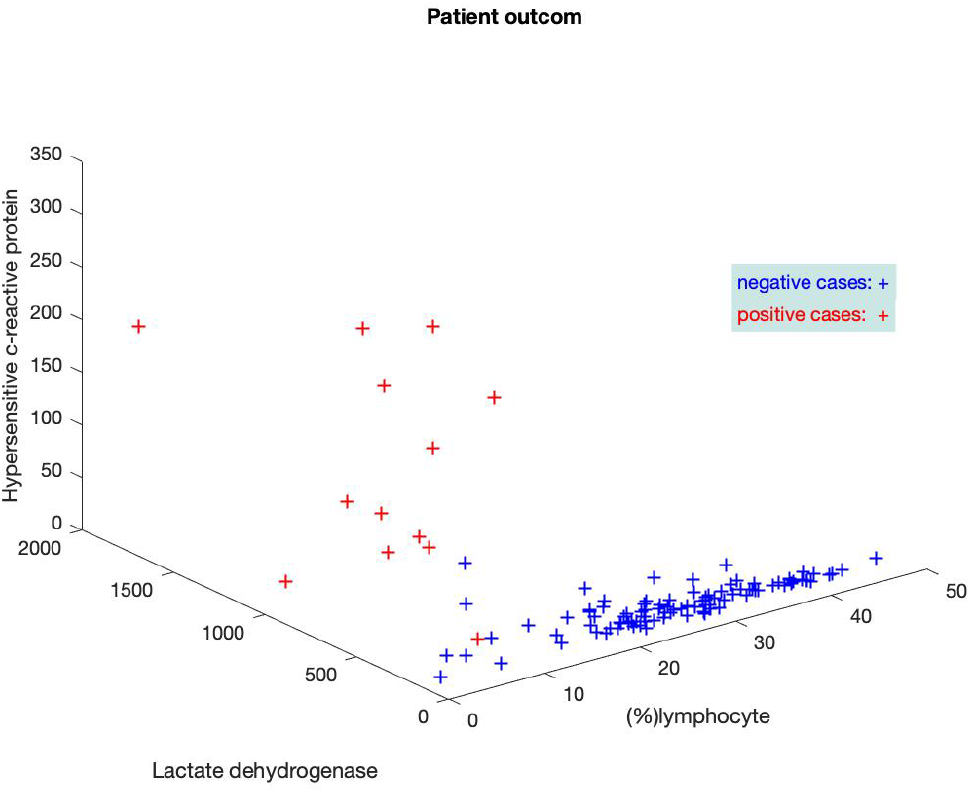
Patient outcome in *test_set*

## 3 Feature analysis

In [1] feature analysis resulted in determination of three key features (lactic dehydrogenase, lymphocytes, and high-sensitivity c-reactive protein) out of 10 most promising features found with optimal XGBoost classifier output. Here, feature analysis is carried out in two steps: 1) a Kohonen neural network (*SOM*) is used for transforming the feature data into component planes *CP*, and 2) a Greedy algorithm is used for finding the minimum dominant set *MDS* of features based on their mutual correlation. Both, *CP* and *MDS* can be rendered and visually inspected. Figure 3 shows the component maps of 10 features selected by [1] after training *SOM* with 344 complete data samples in *train_data*. The map size of SOM was 8 · 3 = 24 neurons.

**Fig. 3.**
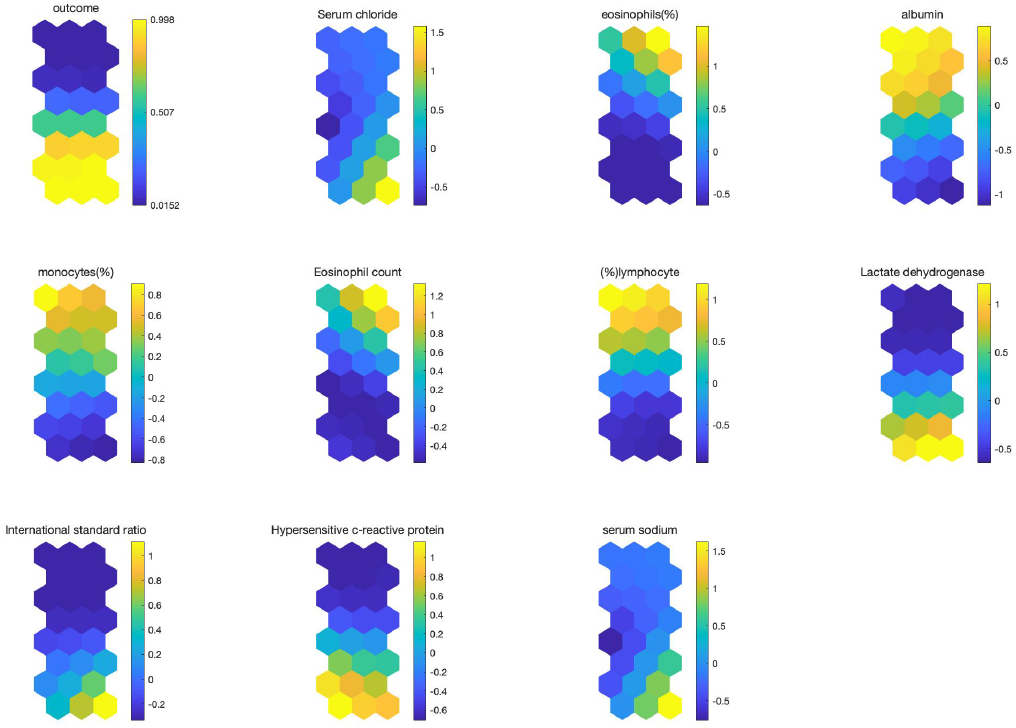
Component planes *CP* (map size [8, 3]) of 10 features and (patient) outcom created by training *SOM* network with *train_data*

The component planes represent the weights of the respective feature in each neuron (hexagon) of the *SOM* map. Each map position (hexagon) represents the weight value in color, and together with its neighbours around it corresponds with similar feature vectors of the training set. A good approach for analyzing is to look for boundaries and color changes in a component plane and similar situations in other planes (colors must not be similar). In this way we recognize good matches in Figure 3 between patient outcome, Lactate dehydrogenase, %lymphozyte, hypersensitive c-reactive protein, and with some restrictions also albumin, and International standard ratio. The first three features correspond to the key features selected for *decTree* and replicate the results of XGBoost classifier.

The second step of our feature analysis is to find the minimum dominant set *MDS* of features. For this, the mutual correlation of feature elements are evaluated and a greedy algorithm searches *MDS* after determination a threshold for mutual feature correlation [7]. Figure 4 shows the resulting correlation graph. We see the key features cover very well the *MDS* when we restrict it to the features with good matches with patient outcome. A strictly reduced *MDS* would consist only of %lymphozyte and hypersensitive c-reactive protein,

**Fig. 4.**
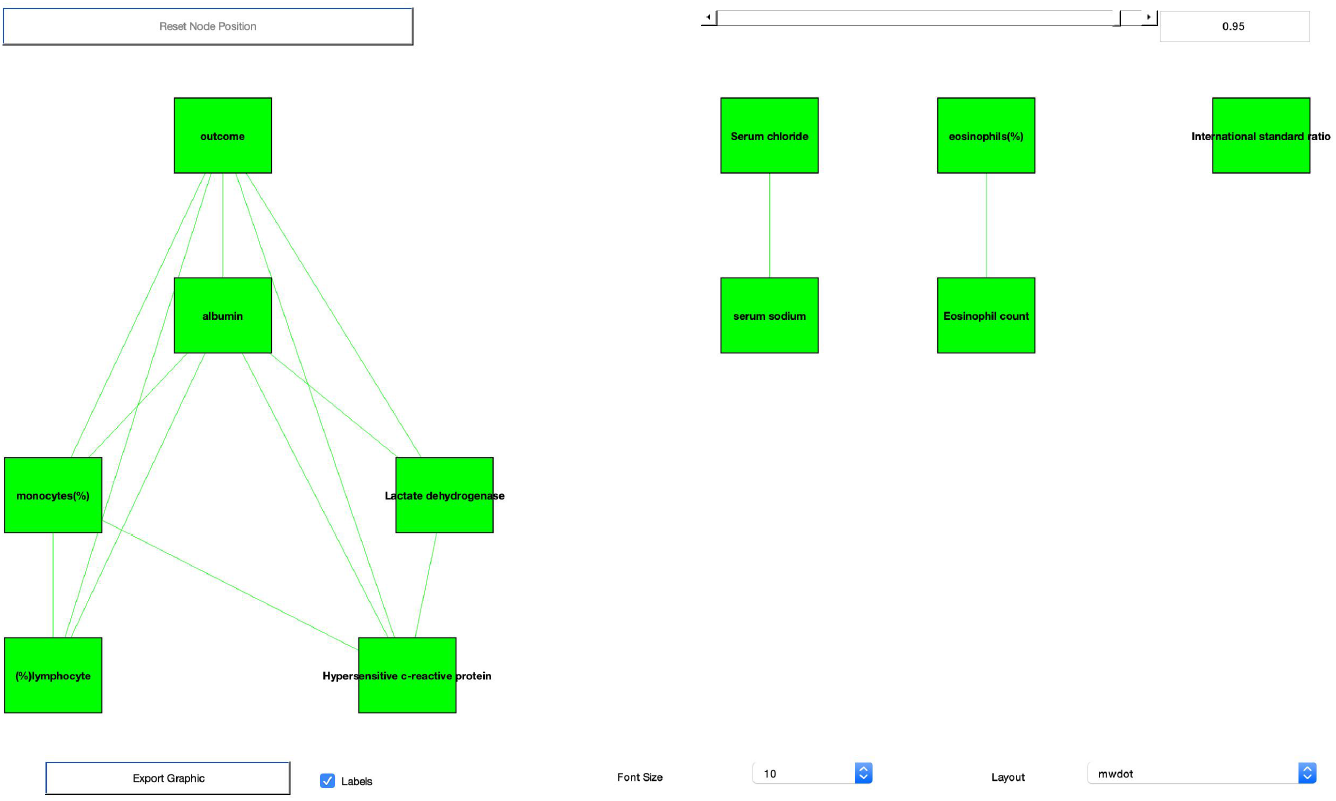
Correlation graph (lines show mutual correlation with threshold = 0.95)

## 4 Building a prediction model

[1] used a decision tree *decTree* to predict patients at the highest risk. Even if the classifier performance is good enough to be used in a operable environment the binary output of *decTree* conceals the grade to which a patient’s feature vector belongs to one of the classes *positive* or *negative*. I assume a human decision maker would prefer to refer to the technical estimation of risk grades when finally deciding about the risk and clinical treatment of patients. Furthermore, the mapping of feature elements by humans likely will rather be in terms like *small, high*, or something unsharp like that than in sharply defined intervals. Fuzzy systems enable the description of models based on Fuzzy rules of type *R*_*i*_ : *IFx*_1_ *is small AND x*_2_ *is large* … *THEN y*_*i*_ *is small* with input vector *X* = (*x*1, *x*2,…)^*t*^ in the premise (*IF*) part and output *y*_*i*_ in the conclusion (*THEN*) part. Building up a Fuzzy model requires first the definition of unsharp terms like *small, medium*,…, so called Fuzzy terms, covering the input elements (*fuzzyfication*) and second the generation of the rule base *R* = *{R*_*i*_ | *i* = 1, 2, …, *N}* describing the complete mapping of the input space into the output function. Finally, the mapping of the rule’s outputs *y*_*i*_ (*accumulation and defuzzyfication*) into a sharp output value *y* = *f* (*y*_*i*_| *i* : 1, …, *N*), *y∈* R has to be established. Fortunately, there are a lot of machine learning tools that can automatically generate an operable Fuzzy model from a training data set (supervised learning).

I used a Sugeno-type Fuzzy modell (there is no need for *defuzzyfication*) and Matlab© function *ANFIS* for generating and training of the Fuzzy model. With *train_data* and three key features as input *ANFIS* creates a Fuzzy model *FIS* with three Fuzzy terms per input (feature) element and *N* = 3^3^ = 27 rules. The model is trained by *ANFIS* with 10 epochs and *train_data* with all 351 patient’s final data samples only. Figure 5 shows prediction results of *FIS* for validation with *train_data*. Figure 6 shows the results of classifier *FIS* with external test data *test_data* (patient’s final data samples only). The performance data of this *FIS* classifier are also displayed in Table 1 (column *FIS_3*) in section 5.

**Table 1.** Performances of *FIS* classifiers for validation with *train_data* and testing with *test_data*

| **FIS model** | *FIS_3* |  |  | *FIS_5* | *FIS_trend* |  |
| --- | --- | --- | --- | --- | --- | --- |
| validation /<br>test data | *train_data* | *train_data* / 10-fold<br>cross validation | *test_data* | *train_data* | *train_data* | *test_data* |
| final samples | 351 | 315/35 | 110 | 344 | 195 | 79 |
| errors $E$ | 5 | $2.6 \pm 2.3$ | 3 | 4 | 0 | 4 |
| $AUC$ | 0.986 | $0.92 \pm 0.002$ | 0.951 | 0.988 | 1 | 0.868 |
| precision $Pre$ | 0.981 | $0.946 \pm 0.008$ | 0.857 | 0.987 | 1 | 0.9 |
| recall $Rec$ | 0.987 | $0.887 \pm 0.003$ | 0.923 | 0.987 | 1 | 0.75 |
| accuracy $Acc$ | 0.986 | $0.926 \pm 0.002$ | 0.973 | 0.988 | 1 | 0.949 |

**Fig. 5.**
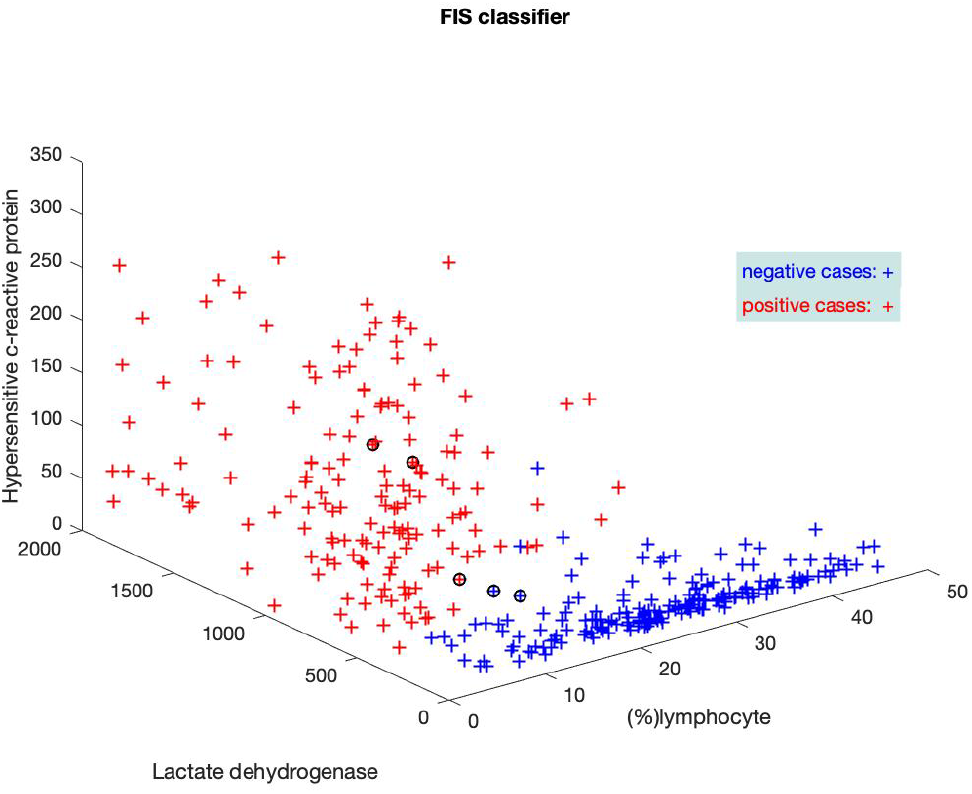
Prediction results of FIS classifier validated with *train_data* (misclassified elements circled)

**Fig. 6.**
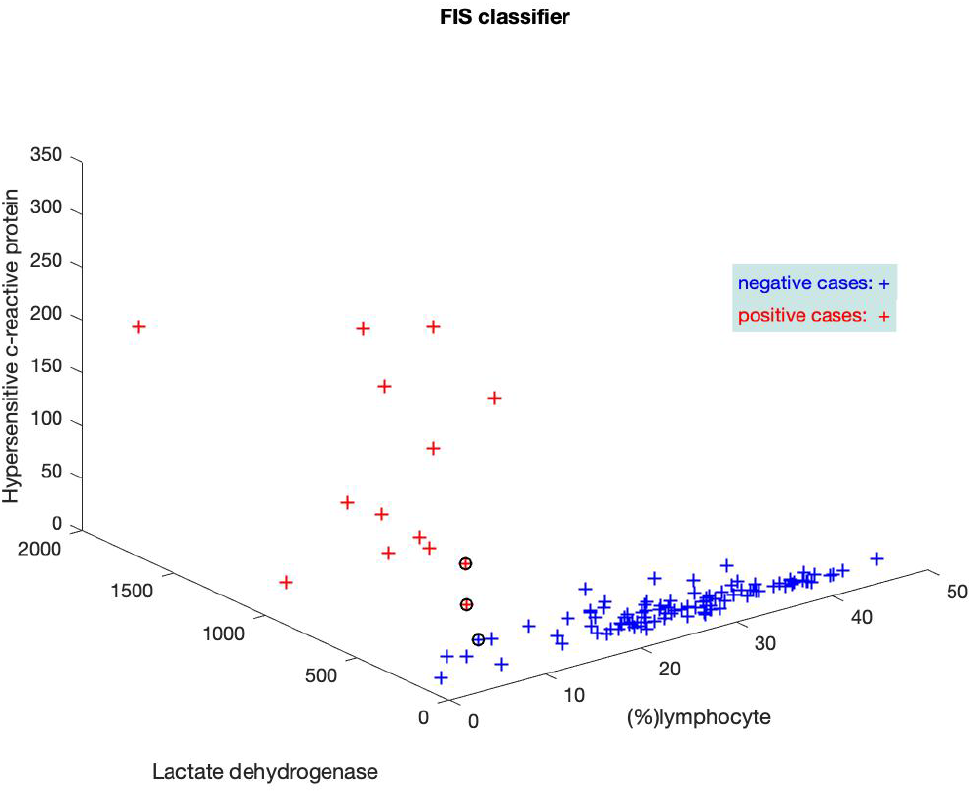
Prediction results of FIS classifier tested with *test_data* (misclassified elements circled)

## 5 Estimation of prediction output

There are various possibilities for improvement of the *FIS* classifier. In general, a Fuzzy system enables to create any nonlinear function or mapping of the input space. From a system perspective one can increase the number of Fuzzy terms for the input and/or output variables thus enlarging the rule base and getting a more detailed function approximation, but at the risk of model overfitting. From a problem perspective one can consider an increased number of input variables - if not already optimally chosen by feature analysis. In the latter perspective I analysed two approaches: 1) additional features (biomarkes) in the input vector, and 2) trend of key features over time. For evaluation of the classifier results the following performances measures have been considered based on [1]: total number of classification errors *E, AUC* score of ROC curve, precision *Pre* = *TP/*(*TP* + *FP*), recall *Rec* = *TP/*(*TP* + *FN*), and accuracy *Acc* = (*TP* + *TN*)*/*(*TP* + *TN* + *FP* + *FN*); TP, TN, FP and FN stand for true positive, true negative, false positive and false negative rates, respectively.

As mentioned in section 3 there are two other features with good resemblance to patient outcome: albumin and International standard ratio. I trained a *FIS* classifier with now five feature elements. *train_data* contains in total 344 complete final data samples with the selected five features. Validation results with this *FIS* classifier are displayed in column *FIS_5* in Table 1. We see a slight improvement in all performance measures. Unfortunately *test_data* doesn’t contain these additional features and therefore a test with the external test data was not possible.

Even as a medical layperson, it can be assumed that the patient’s physiological state and the health risk can also be judged by the development of the biomarkers and not only by their last value. Therefore, I tried to include the biomarkers’ trend in time into the model for risk assessment. However, it is now the case here that the blood samples and thus the biomarkers in the data records were not systematically recorded over time. Thus, it is only possible to determine the temporal trend here as an example. To do this, I simply chose the difference between the last and penultimate data sample as a measure for the trend over time. The *FIS* model *FIS_3* with the three key features is used as the model basis and the input vector is expanded with the features’ trend values.Validation and test results with this *FIS* classifier are displayed in column *FIS_trend* in Table 1. These results were achieved regardless of whether the trend of one, two or all three key features was used in the *FIS* model *FIS_trend*. Since the blood values were obviously not systematically recorded, only 195 trend values could be determined in *train_data* for training and validation, and only 79 in *test_data* for testing the classifier. And thus the results are not similarly representative like those of the other models *FIS_3* and *FIS_5*.

## 6 Discussion

This study shows the potential of Fuzzy models for the risk assessment of COVID-19 patients in several ways. First of all, the results in [1] could be replicated and at the same time an improvement of the risk assessment could be achieved. The consideration of the temporal development of the biomarkers in the models had a decisive influence on the model performance. However, this could not be tested in detail because the training and external test data contained too few examples and in particular the blood samples had not been recorded systematically over time.

In addition, a non-binary risk assessment has been introduced. This supports an interpretation of the model output by medical professionals. If one looks at the real risk values that model *FIS_3* calculates for the external test data, one can see that the wrongly classified items in most cases are very close to the decision limit 0.5 for binarization. Figures 7 and 8 show the statistical values of the risk assessment with the training and test data using boxplots. After assigning the incorrectly classified examples (FN = 2, FP = 1 in Figure 8), the associated boxplot shows that the real output values are close to the decision value 0.5 and can therefore be better assessed by a medical expert than the wrong binary decision. The continuous model output enables further opportunities for technical support for medical experts in COVID-19 risk assessment.

**Fig. 7.**
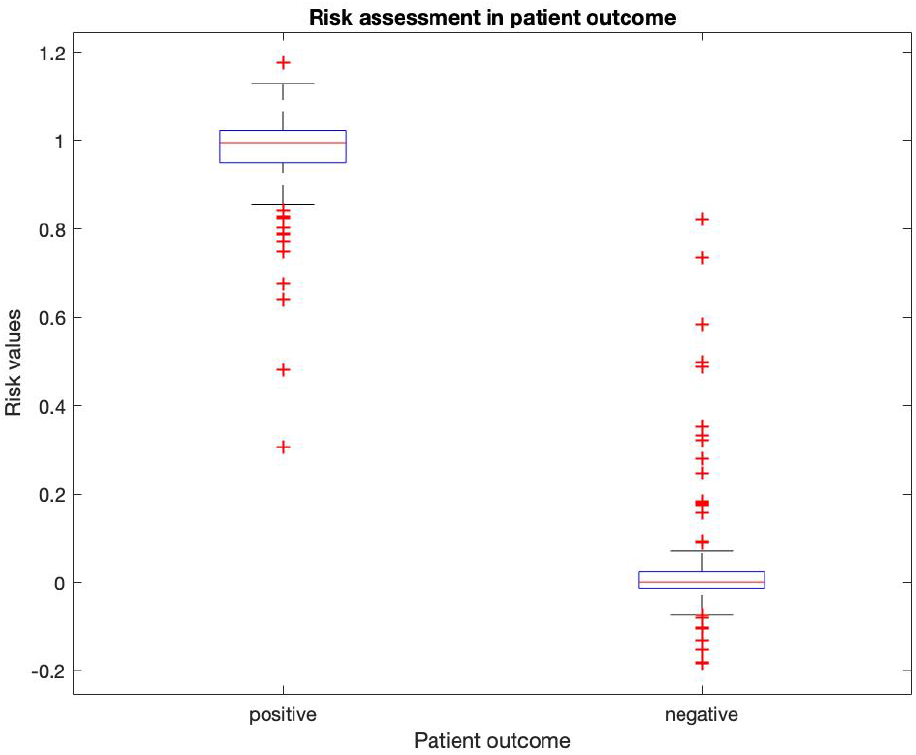
Statistical distributuion of risk assessment (real model outputs) with *FIS_3* Fuzzy model and training data *train_data*, boxplots with 25, 75 percentil (blue), median and outliers (red)

**Fig. 8.**
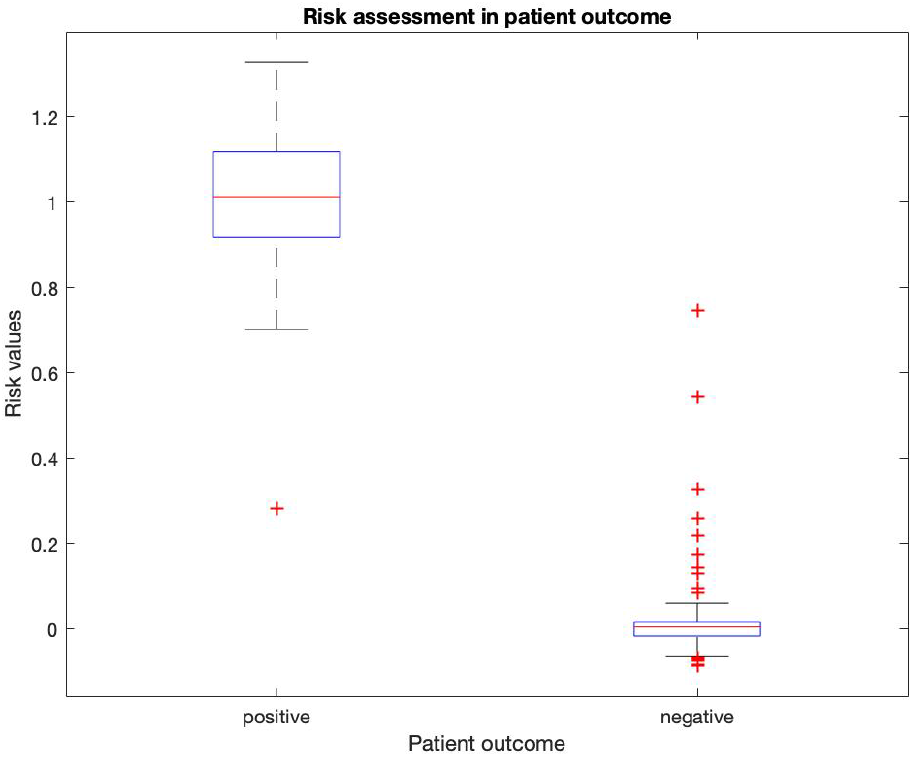
Statistical distributuion of risk assessment (real model outputs) with *FIS_3* Fuzzy model and training data *test_data*, boxplots with 25, 75 percentil (blue), median and outliers (red)

## 7 Conclusion

In summary, this study introduces a Fuzzy logic based prediction system for COVID-19 risk assessment with improved performance compared with other approaches in literature ([1]). This provides a good basis for the development of a transparent and operational system for risk assessment of COVID-19 patients. In addition to the selection and consideration of the final biomarker or feature elements, a model extension is also successfully tested that takes their changes over time into account. The model output is non-binary and is therefore particularly suitable for a decisive interpretation by medical experts. A further investigation of the time horizon of the risk assessment was initially not carried out since the blood samples were not recorded systematically in the currently available training and test data.

## Data Availability

Basically, all data for feature analysis as well as for training and testing the classifiers have been taken from the original data base provided by L. Yan et al.

## Acknowledgements

I would like to thank Jorge Goncalves and the authors in [1] for providing the training and test data, and in particular Jorge for sharing and discussing this topic with me.

